# Immunogenicity and crossreactivity of antibodies to SARS-CoV-2 nucleocapsid protein

**DOI:** 10.1101/2020.12.19.20248551

**Authors:** Carlota Dobaño, Rebeca Santano, Alfons Jiménez, Marta Vidal, Jordi Chi, Natalia Rodrigo Melero, Matija Popovic, Rubén López-Aladid, Laia Fernández-Barat, Marta Tortajada, Francisco Carmona-Torre, Gabriel Reina, Antoni Torres, Alfredo Mayor, Carlo Carolis, Alberto L. García-Basteiro, Ruth Aguilar, Gemma Moncunill, Luis Izquierdo

**Author notes:** Contributed equally (Gemma Moncunill, Luis Izquierdo). **Conflict of interest statement** The authors have declared that no conflict of interest exists.

## Abstract

COVID-19 patients elicit strong responses to the nucleocapsid (N) protein of SARS-CoV-2 but binding antibodies are also detected in prepandemic individuals, indicating potential crossreactivity with common cold human coronaviruses (HCoV) and questioning its utility in seroprevalence studies. We investigated the immunogenicity of the full-length and shorter fragments of the SARS-CoV-2 N protein, and the crossreactivity of antibodies with HCoV. We indentified a C-terminus region in SARS-CoV2 N of minimal sequence homology with HCoV that was more specific and highly immunogenic. IgGs to the full-length SARS-CoV-2 N also recognised N229E N, and IgGs to HKU1 N recognised SARS-CoV-2 N. Crossreactivity with SARS-CoV-2 was stronger for alpha-rather than beta-HCoV despite having less sequence identity, revealing the importance of conformational recognition. Higher preexisting IgG to OC43 N correlated with lower IgG to SARS-CoV-2 in rRT-PCR negative individuals, reflecting less exposure and indicating a potential protective association. Antibodies to SARS-CoV-2 N were higher in patients with more severe and longer symptoms and in females. IgGs remained stable for at least 3 months, while IgAs and IgMs declined faster. In conclusion, N is a primary target of SARS-CoV-2-specific and HCoV crossreactive antibodies, both of which may affect the acquisition of immunity to COVID-19.

## INTRODUCTION

The identification of the antigens and epitopes that induce antibody responses after exposure to SARS-CoV-2 infection is one of the requirements to estimate the seroprevalence in a population. In addition, it is also essential to understand immunity to COVID-19 disease. Previous knowledge on other related coronaviruses and the prompt sequencing of the SARS-CoV-2 genome early in the pandemic allowed to identify the spike (S) (1) and the nucleocapsid (N) (2) structural proteins as major targets of antibodies. Consequently, both antigens constituted the basis for most immunoassays developed to study COVID-19 distribution and protective immune responses. The surface glycoprotein S, which contains the receptor binding domain (RBD), has a better known function in immunity (3–5) and is the leading antigen candidate for vaccine development (6, 7). N is smaller than S, lacks a glycosylation site, and is extensively used in leading serodiagnostics kits (8–11) due to its abundant expression during infection (12–14) and early antibody response (15, 16), but its immunological relevance is less established. The pattern of antibody responses to S compared to N may vary according to disease severity (17) and age (18). N forms ribonucleoprotein complexes during the virion assembly process by binding to the viral RNA genome and packing it into long helical structures (19). Its main function is to regulate viral RNA transcription during replication, promoting the synthesis of its own proteins (14) while interfering with the metabolism (19, 20), protein translation (21) and proliferation (22) of the infected host cell. During the process of infection, N dissociates itself from the genome and is exposed to the host immune system (23), and its high immunogenicity has also prompted its exploration as vaccine target (13, 24).

We recently developed a multiplex quantitative suspension array assay using the xMAP Luminex technology (25). We included antigen fragments of S, N and membrane (M) SARS-CoV-2 proteins to establish the utility of each construct for seroprevalence and correlates of immunity studies. Initial investigations using plasma or serum samples from individuals infected with SARS-CoV-2 and prepandemic samples (negative controls), showed different levels of immunogenicity and specificity across the antigens and fragments tested. Even though antibodies to full-length N (N FL) constructs were usually high in adult COVID-19 cases, moderate to high responses were frequently detected in samples collected before the pandemic (25), as it had been reported before also for SARS-CoV-1 samples (26). This questioned the use of N FL for seropositivity calculations that were consequently restricted to S-based antigens in our initial evaluations (27). We hypothesized that such antibody signals in negative controls, more prominent for N than S antigens, reflected crossreactivity with human coronaviruses of the common cold (HCoV) due to highly conserved regions, rather than nonspecific or polyreactive responses. The HCoV include alphacoronaviruses (229E and NL63) and betacoronaviruses (HKU1 and OC43). In fact, increasing number of reports in the literature are consistent with some antibodies to HCoV being crossreactive with the betacoronaviruses SARS-CoV-2 and SARS-CoV-1 (28–32). Interestingly, significant protein similarity between SARS-CoV-1, SARS-CoV-2 and other HCoV has been reported for N, including a highly conserved motif in the N-terminal (NT) half of the protein (FYYLGTGP) (33) and relevant immunodominant epitope regions (24, 34). Likewise, preexisting SARS-CoV-2-specific T cells have also been reported and attributed to crossreactivity with HCoV previously encountered (35–39).

A key question is the relevance of those preexisting antibodies on acquisition of COVID-19 immunity. Is this crossreactivity sufficient to protect against disease? (40). If this is the case, it could be one of the reasons why children may be more protected than older adults (41) (42, 43). Alternatively, these preexisting crossreactive antibodies could interfere with the development and/or maintenance of effective levels of SARS-CoV-2 antibodies (44, 45) and, even worse, they could have a negative impact by mediating antibody-dependent disease enhancement (ADE) (46, 47), which could be associated with severe prognosis (48, 49). Our study aimed to better characterize the immunogenicity, specificity and crossreactivity of anti-N antibodies from SARS-CoV-2 and 229E, HKU1, NL63 and OC43 HCoV, measured simultaneosuly. To this end, we tested different antigenic fragments in multiplex and different Ig isotypes, and compared their relative immunogenicity in prepandemic and pandemic samples, including SARS-CoV-2 positive cases. In addition, we aimed to better understand the demographic, clinical and epidemiological variables affecting the levels of antibodies to N SARS-CoV-2 in exposed people. Our study helps addressing the extent and characteristics of this crossreactivity in the immune response to COVID-19 and the utility of various N-based antigens in serodiagnostics and seroprevalence studies.

## RESULTS

### Immunogenicity, specificity and seropositivity of SARS-CoV-2 N antibodies

Plasmas from individuals diagnosed with COVID-19 had statistically significantly higher levels of IgG, IgA and IgM to the FL as well as the NT and C-terminal (CT) domains of SARS-CoV-2 N protein (**Figure S1**) than prepandemic plasmas (**Figure 1**). Levels were statistically significantly higher in patients who were hospitalized compared to non-hospitalized (asymptomatics and mild cases) (**Figure 1A**). However, prepandemic plasmas also contained IgG, IgM and IgA antibodies recognizing N antigens. In the case of IgG, the responses against N FL were the highest, followed by the N NT and N CT domains. This high reactivity of negative control plasmas resulted in a higher seropositivity cutoff and lower seroprevalence estimates for IgG to N FL in our cohort of health care workers, with 10.1% at M1 (**Figure S2A**) compared with 11.3% for RBD (27). Higher plasma dilutions (1/3500 vs. 1/500) reduced antibody levels proportionally more in prepandemic than pandemic samples, increasing signal-to-noise ratio, sensitivity and overall seropositivity (**Figure S2B**).

**Figure 1.**
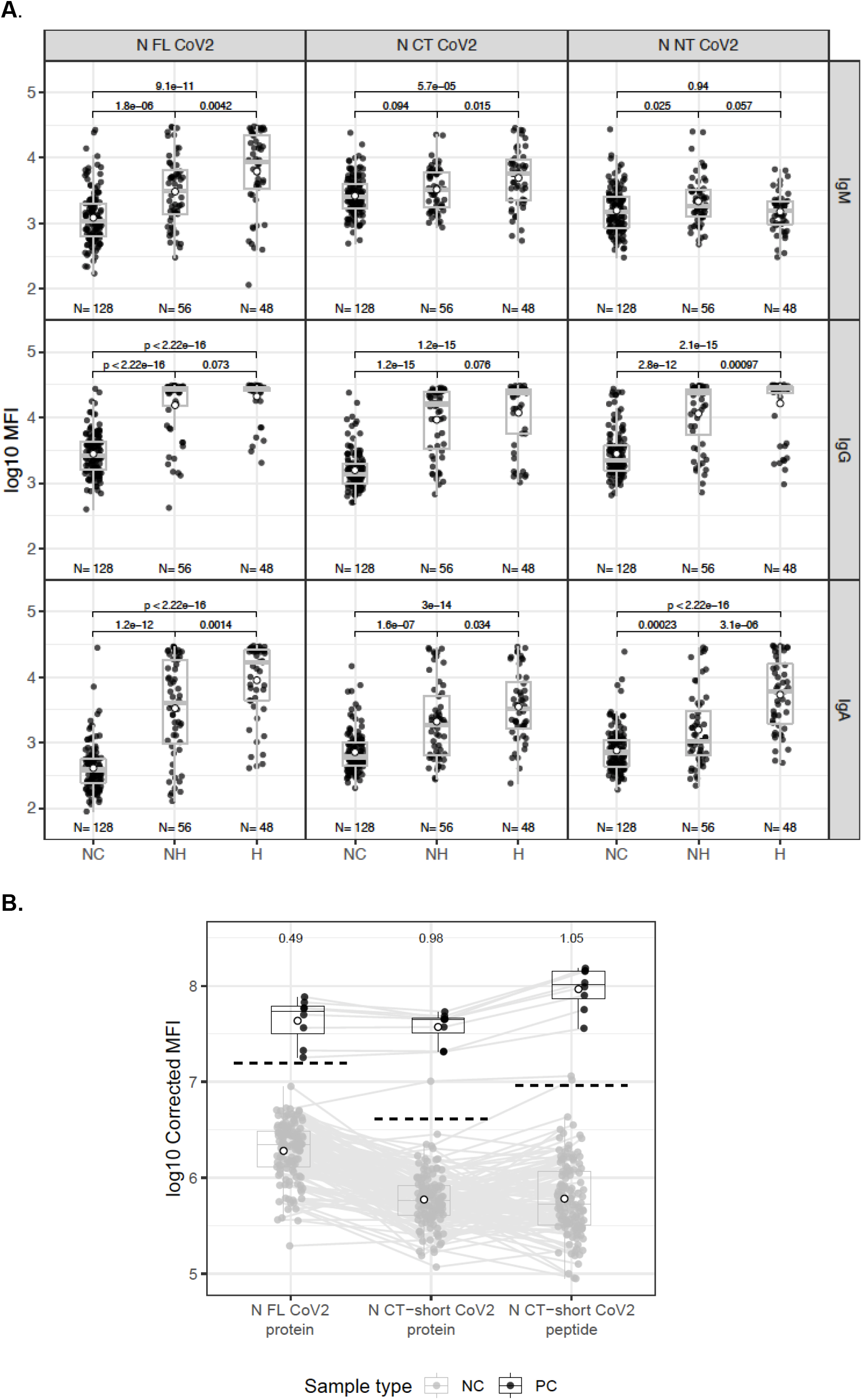
Levels (log_10_ median fluorescence intensity, MFI) of antibodies to nucleocapsid (N) SARS-CoV-2 antigens in prepandemic (NC) and pandemic samples. The lower and upper hinges of the boxplots correspond to the 25^th^ and 75^th^ percentiles (IQR) and extend 1.5 * IQR from the hinge; open circles are means. **A)** N FL: full-length produced at ISGlobal; N C-terminus [CT] and N N-terminus [NT] domains produced at CRG. NH: non-hospitalized, H: hospitalized. P values were calculated by the Wilcoxon test. **B)** N FL and N CT-short from ISGlobal and N CT-peptide. Numerical values on the top of the boxplots are the ratios of the means of positive controls (PC) vs. seropositive cutoff values indicated by black dashed lines, calculated as 10 to the mean plus 3 standard deviations of the log_10_-transformed MFI of the NC. The grey lines link the samples from the same individuals. PC are the different points of the titration curve corrected to have the same dilution factor.

Nevertheless, seroreactivity in prepandemic samples was still patent, which could be attributed to the presence in our antigens of stretches of immunogenic aminoacid sequences similar to other HCoV (24, 28). Searching for SARS-CoV-2 specific responses, we tested a CT-short protein and a CT-peptide from N that had a lower percentage of identity with HCoVs N (**Figure S1**). The shorter N CT constructs were immunogenic in pandemic positive samples and less seroreactive in prepandemic samples than N FL, with higher signal-to-noise ratios obtained (**Figure 1B**). Additional information on the serological characterization of the rest of N FL constructs is provided in the **Supplemental material, Figures S3** & **S4**.

### Crossreactivity of IgG antibodies to SARS-CoV-2 and HCoV N FL proteins

To test whether the antibodies recognizing SARS-CoV-2 N antigens in negative samples could be due to crossreactivity with HKU1, OC43, NL63 or 229E HCoV, we compared the patterns of IgG responses to N FL from the five coronaviruses in prepandemic and COVID-19 samples at two timepoints, also including the SARS-CoV-2 N CT-short fragment. Overall, anti-N IgG levels to SARS-CoV-2 were significantly higher in COVID-19 cases vs. prepandemic samples compared to HCoV IgG levels, which were similar or slightly higher in pandemic positive vs. prepandemic samples, like for NL63 (**Figure 2A**). This pattern indicates both high immunogenicity of SARS-CoV-2 N antigens in COVID-19 samples, but also some level of crossreactivity against some HCoV N antigens. Furthermore, data support the higher SARS-CoV-2-specificity of the IgG response to the CT-short portion of N because IgG levels in prepandemic samples were the lowest and similar to pandemic real time reverse-transcriptase polymerase chain reaction (rRT-PCR) negative (**Figure 2A**). Pandemic samples from rRT-PCR negative health care workers had significantly higher levels of IgG to N FL, but not N CT-short, than prepandemic samples and significantly lower levels than rRT-PCR positive individuals.

**Figure 2.**
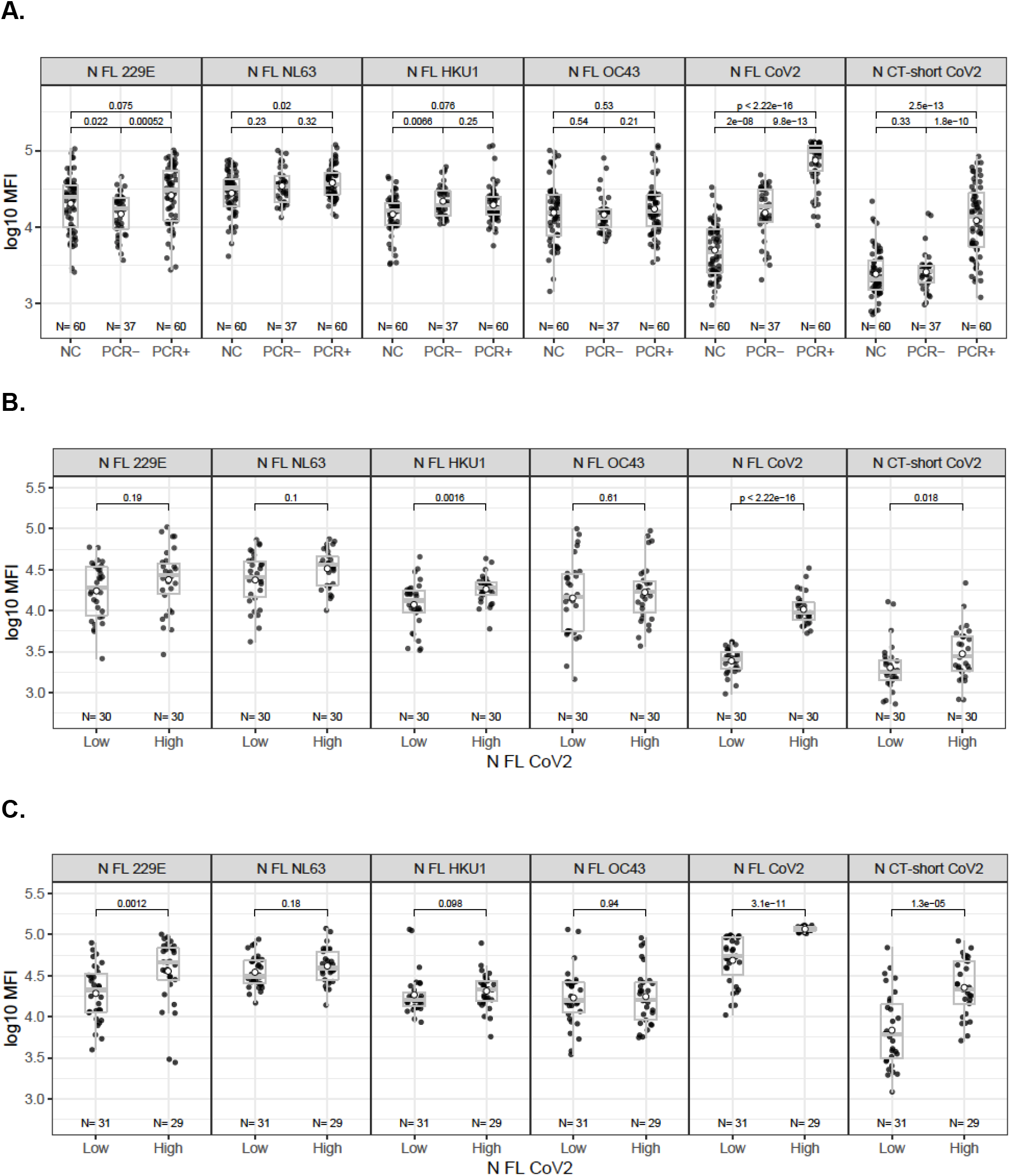
Levels (log_10_ median fluorescence intensity, MFI) of IgG antibodies to N antigens of human coronaviruses. **A)** Plasmas from prepandemic (NC) and pandemic (rRT-PCR positive vs. negative) individuals. Plasmas stratified according to low vs. high IgG responses to SARS-CoV-2 N FL in **B)** prepandemic and **C)** pandemic rRT-PCR positive. Boxplots indicate median and IQR, open circles are means, and p values were calculated by Wilcoxon test.

In prepandemic samples, IgG levels to HKU1 HCoV N FL were statistically significantly higher in samples with higher IgG levels to SARS-CoV-2 N FL (**Figure 2B**), and there was a significant correlation (rho=0.35, p=0.0065) between N FL IgG responses of HKU1 HCoV to SARS-CoV-2 and to a lesser extent with NL63 HCoV (**Figure 3A**), suggestive of crossreactivity. In pandemic rRT-PCR positive samples, IgG levels to 229E HCoV N FL were statistically significantly higher in samples with higher IgG levels to SARS-CoV-2 N FL (**Figures 2C**), and there was a significant correlation of N FL IgG responses between SARS-CoV-2 and 229E HCoV (rho=0.38-0.45, p<0.0024) (**Figures 3A** & **3B**). Correlation between N FL and N CT-short was higher and more significant in pandemic (rho=0.79, p<0.001) and rRT-PCR positive (rho=0.6, p<0.001), compared to prepandemic and rRT-PCR negative samples, respectively (**Figure 3A** & **3B**). In contrast, there was a significant inverse correlation between OC43 HCoV and SARS-CoV-2 IgG levels in rRT-PCR negative pandemic samples (rho=-0.39, p=0.019) (**Figure 3B**). Integrating the magnitude of responses to the HCoV N FL proteins, the correlation with SARS-CoV-2 N FL was statistically significant in pandemic (rho=0.29, p=0.026, **Figure 3B**) and rRT-PCR positive samples (rho=0.27, p=0.0084, **Figure 3C**), and it was higher and only significant for alpha-(229E and NL63) than for beta-HCoV (229E and NL63) (**Figure 3C**). The correlation in prepandemic samples improved when OC43 beta-HCoV was excluded from the HCoV summation (rho=0.25, p=0.053 vs. rho=0.2 p=0.13), although IgG levels to OC43 strongly correlated with those of alpha-coronaviruses (rho=0.522 for 229E, rho=0.503 for NL63) (**Figure S5A**). SARS-CoV-2 N FL levels correlated more significantly with N NT in prepandemic (rho=0.526) and with N CT in pandemic (rho>0.624) samples. The breadth of responses to human coronaviruses was heterogeneous among individuals, but patterns of antibodies to alpha- and beta-HCoV tended to cluster together (**Figure S5B**).

**Figure 3.**
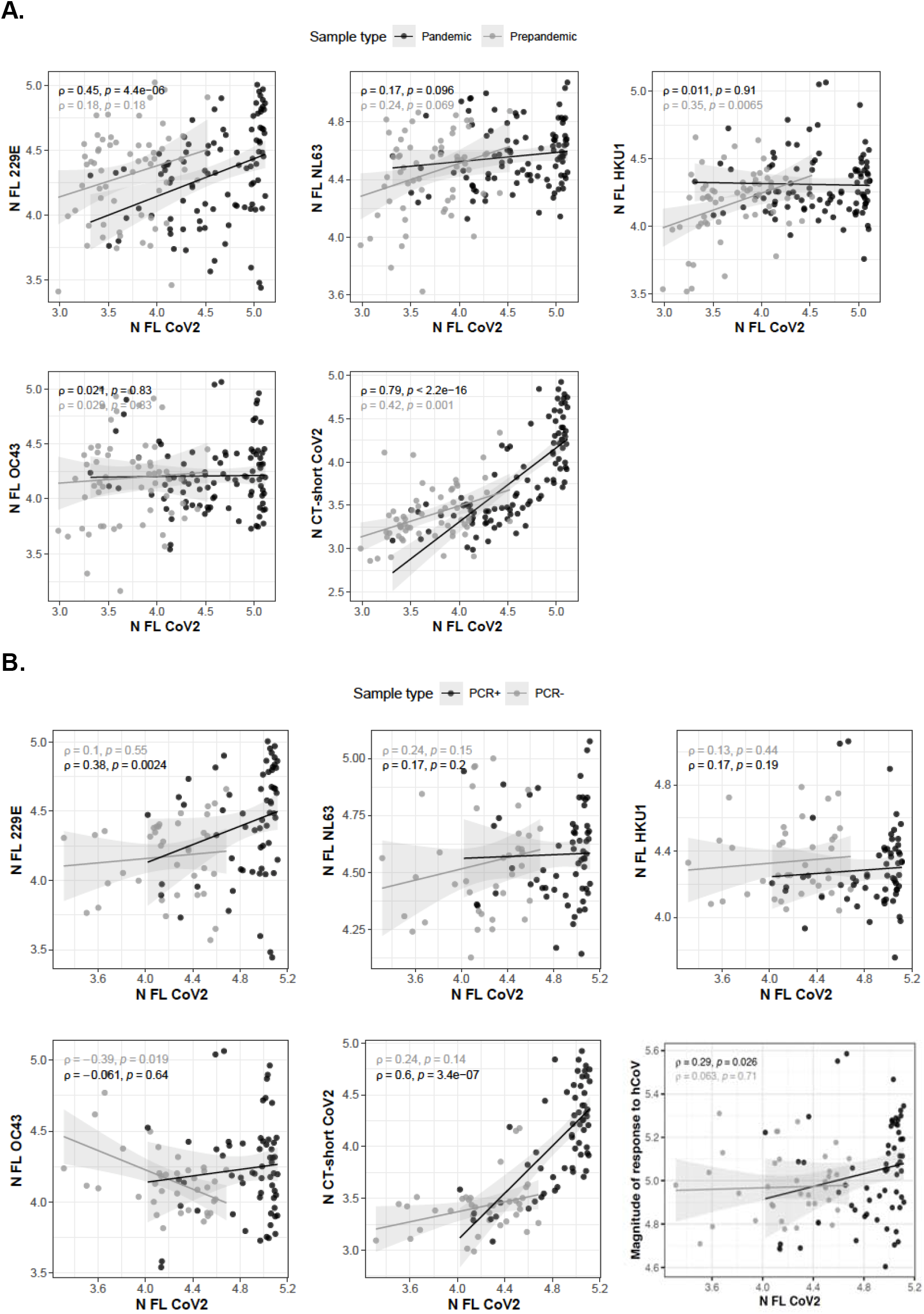
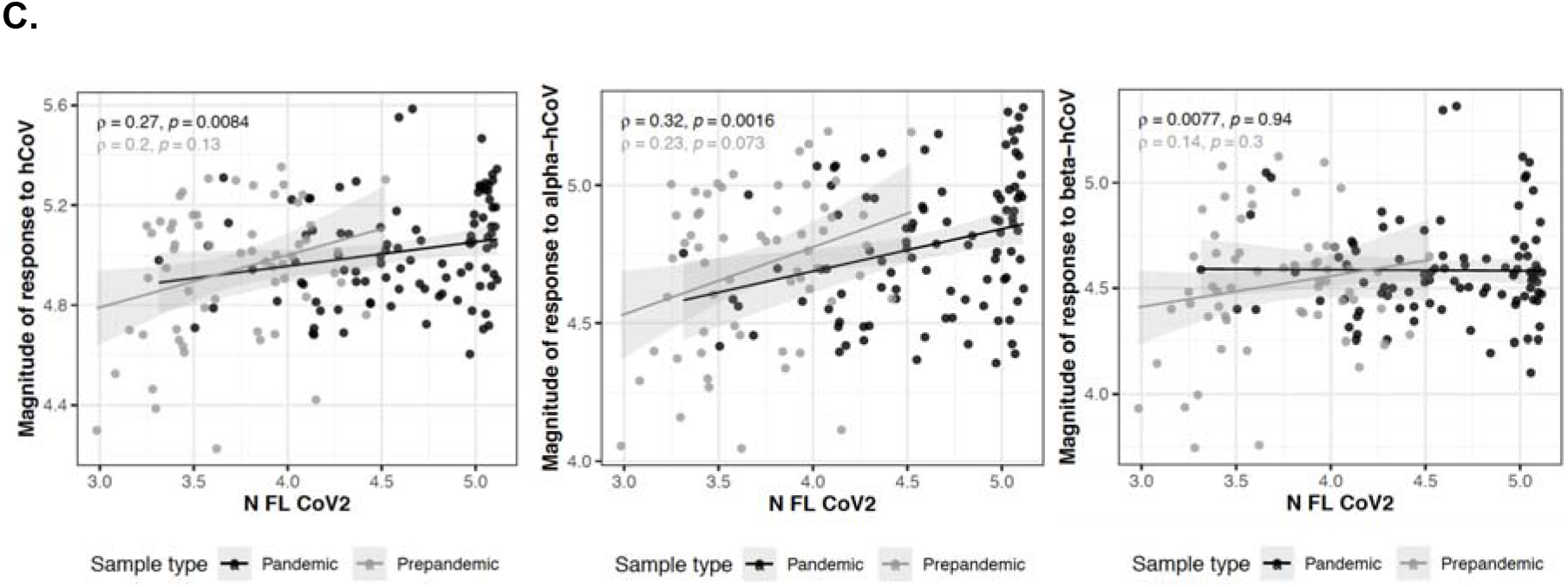
Correlations of plasma IgG levels (log_10_ MFI) to SARS-CoV-2 and HCoV N antigens. **A)** Pandemic and prepandemic samples. **B)** Pandemic rRT-PCR positive and negative. **C)** Magnitude of response to HCoV in pandemic and prepandemic samples. Rho and p values are calculated by Spearman, shaded areas represent 0.95 confidence intervals.

In individuals who became infected with SARS-CoV-2 from M0 to M1, IgG levels increased significantly for SARS-CoV-2 N FL (p=0.015) and SARS-CoV-2 N CT-short (p=0.031), while antibody profiles for HCoV N proteins were variable (**Figure 4**). IgG to N from beta-HCoV tended to decrease or maintain, whereas IgG to N from alpha-HCoV (NL63 and 299E) increased in about half of individuals but overall not significantly due to low sample size and high variability. There did not seem to be a relationship between IgG levels at baseline and the change in levels at month (M)1. In uninfected individuals, IgG levels declined from M0 to M1, significantly for 4 of 6 N antigens (**Figure 4**).

**Figure 4.**
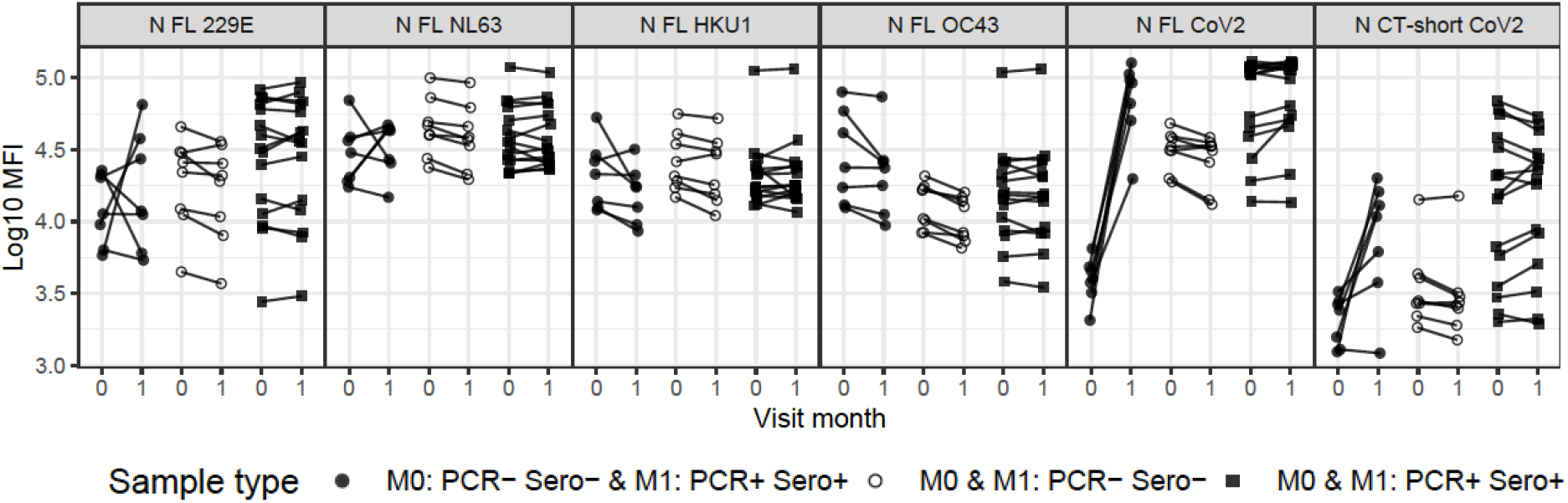
Change in antibody levels to human coronavirus nucleocapsid (N) proteins from baseline (M0) to one month later (M1) depending on the rRT-PCR SARS-CoV-2 infection and seropositivity to RBD status (positive + or negative -).

### Kinetics and determinants of antibody levels to SARS-CoV-2 N FL

Having shown that SARS-CoV-2 infections induce a specific antibody response to the N antigen, despite having in some cases preexisting crossreactive antibodies due to prior exposure to HCoV, we investigated the kinetics and demographic, clinical and epidemiological variables associated with the levels of immunoglobulins to N FL. We analyzed samples from the cohort of health care workers followed up for 3 months in whom we had previously characterized RBD antibodies (27, 50). We performed two analyses, including (i) all individuals and (ii) only those who were seropositive for SARS-CoV-2 RBD antibodies and, therefore, with evidence of exposure to the virus. IgA and IgM to SARS-CoV-2 N FL declined more markedly than IgG, which maintained high levels over the 3 months of study (**Figure 5**).

**Figure 5.**
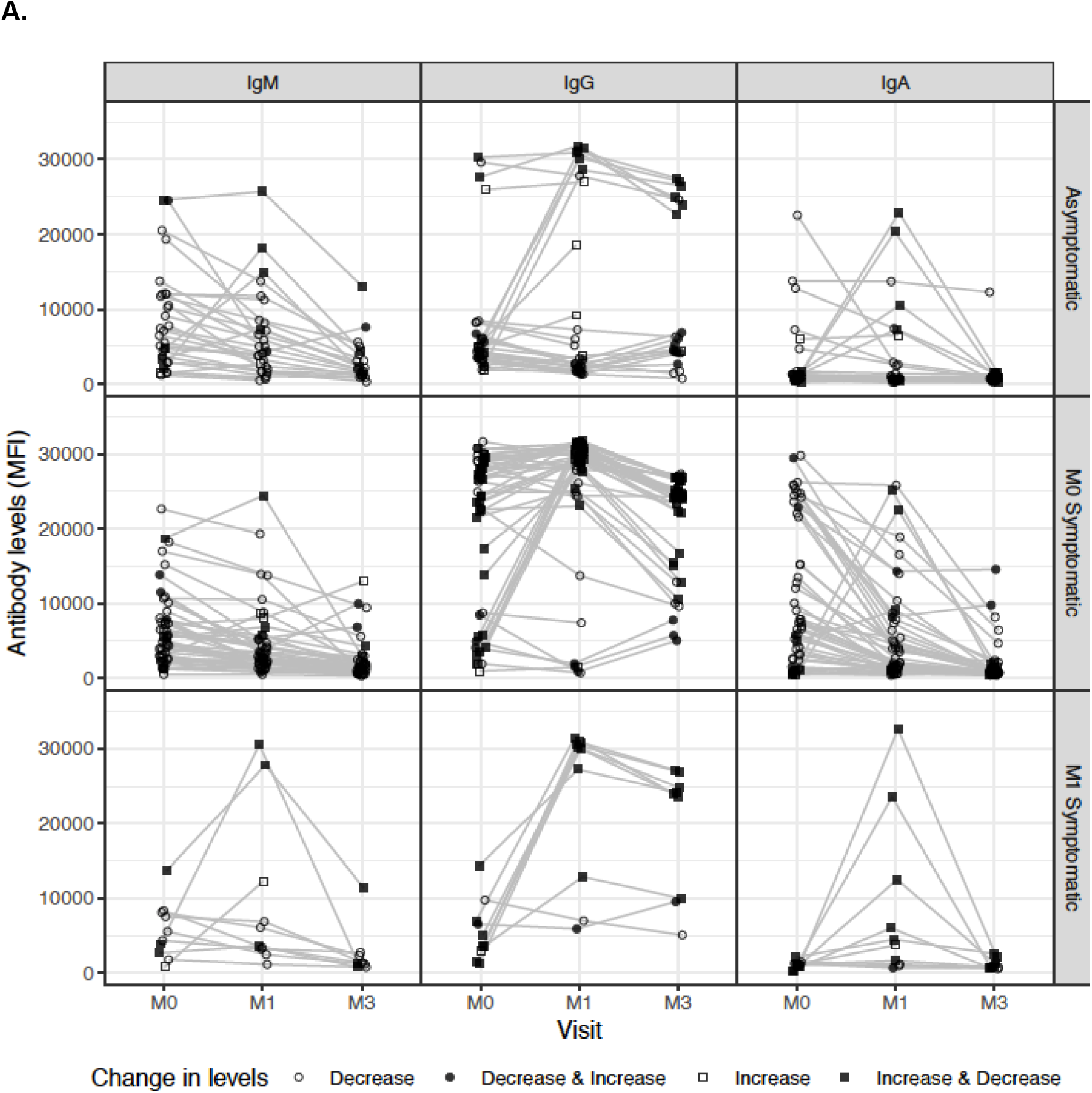
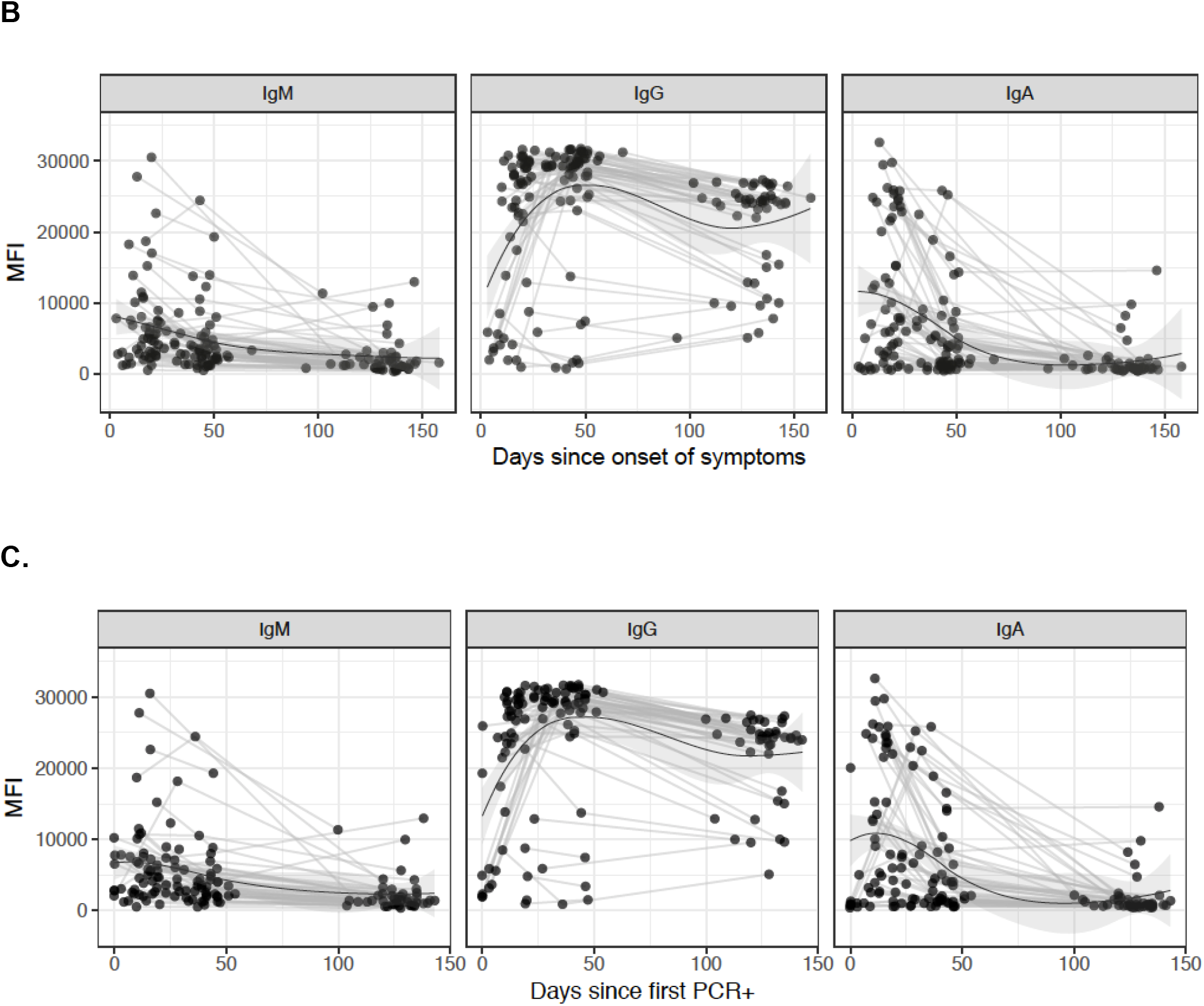
Kinetics of antibody responses to SARS-CoV-2 nucleocapsid (N) full-length protein in a cohort of health care workers with any evidence of infection (rRT-PCR positive or RBD antibody positive for any isotype) at any visit (month [M]0), M1 and M3). **A)** Antibody levels by visit and stratified by symptoms. **B)** Antibody kinetics since onset of symptoms. **C)** Antibody kinetics since first rRT-PCR positive. The curves represent the kinetics of the samples over time and was calculated by the LOESS (locally estimated scatterplot smoothing) method. The shaded area represents the 0.95 confidence intervals.

Having symptoms had a statistically significant association with higher IgA and IgG levels to SARS-CoV-2 N FL, and having a longer duration of symptoms had a statistically significant association with higher IgA and IgM levels to SARS-CoV-2 N FL (**Figure 6A**). IgA levels to SARS-CoV-2 N FL tended to be higher in older individuals, and IgM responses were statistically significantly higher in females than males (**Figure 6B**). No other clear associations were found for occupation or number of COVID-19 contacts.

**Figure 6.**
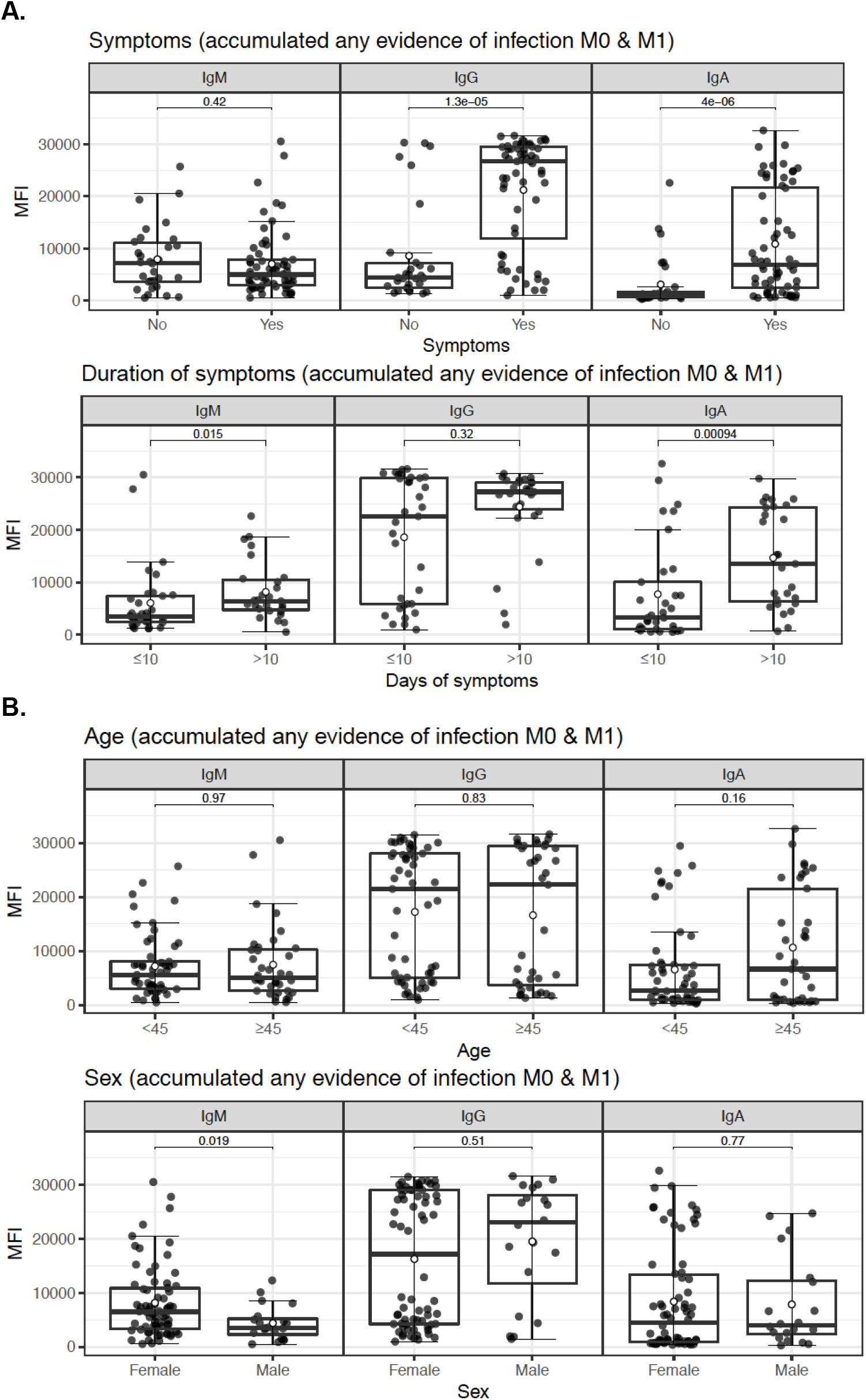
Factors affecting the levels of antibodies to SARS-CoV-2 nucleocapsid (N) full-length antigen. Boxplots indicate median and IQR, open circles are means, and p values were calculated by Wilcoxon test.

## DISCUSSION

Our study shows that SARS-CoV-2 infection induces specific IgM, IgA and IgG antibody responses to epitopes located at both the NT and CT domains of the N protein, consistent with immunogenicity data reported in the literature (15, 20, 51–53). All antibodies were of higher magnitude in patients with more severe clinical manifestations compared to those not requiring hospitalization, and in symptomatic compared to asymptomatic health care workers for IgG and IgA. Longer duration of symptoms was associated with higher IgA and IgM levels, IgA levels tended to increase with age, and females had higher IgM levels.

Importantly, anti-N IgG antibodies remained quite stable over a period of at least 3 months following mild or asymptomatic infection, while IgAs and IgMs declined faster, similar to SARS-CoV-2 RBD responses (50).

We also found that significant levels of IgG, IgM and IgA recognising SARS-CoV-2 N proteins are present in so-called negative controls. This reactivity could limit the utility of the N antigen in seroprevalence studies, as the cutoff values for IgG are higher and hence the percentage of seropositive individuals may be underestimated, compared to calculations based on S antigens (27). However, point prevalence estimates during outbreaks could still be assessed with IgA and IgM to N proteins, as prepandemic samples have lower levels of such preexisting isotypes probably due to their shorter half-life, which is consistent with our kinetics data. The seroreactivity of N in negative controls also leads to lower specificity of SARS-CoV-2 diagnostic tests based on this antigen. To circumvent this, assays with truncated versions of N based on immunogenic regions have been developed (Roche, Abbot) (8, 10), as we have done here with N CT-short and N CT-peptide.

Among the three regions of N (NT RNA-binding domain, central linker, and CT dimerization domain), the NT and CT are the major structural and functional domains (19). Previous works carried out with SARS-CoV-1 N, sharing >90% identity to SARS-CoV-2 N, revealed the presence of four main immunodominant regions (EP1-4) in different domains of the N protein (**Figure S1A**) (24, 28, 54). We focused on the B cell immunodominant domain EP4 showing a lower percentage of aminoacid identity with other HCoVs (residues 356-404) (**Figure S1B**) (55), which corresponds to the CT region of SARS-CoV-2 N. The N CT-short protein and the N CT-peptide presented epitopes that were more specific and still immunogenic, thus applicable for seroprevalence and immunity studies. Correlation between N FL and N CT-short was higher and more significant in pandemic and rRT-PCR positive compared to prepandemic and rRT-PCR negative samples, respectively, indicative of a specific response to SARS-CoV-2 N upon infection.

Interestingly, pandemic rRT-PCR negative individuals had higher responses to N FL than prepandemic samples and significantly lower than rRT-PCR positive individuals. A possible explanation is that some of the rRT-PCR negative individuals might in fact have been exposed to SARS-CoV-2 despite being seronegative for RBD antibodies, but they could have been positive for T cell responses. This could indicate a higher sensitivity of N to detect low-responders or early infections (28) and would imply that a much higher percentage of the population than estimated in the literature could have been exposed to SARS-CoV-2.

Alternatively, the responses in rRT-PCR negatives could be due to crossreactivity with HcoV in this group of health care workers that due to their occupation might be at higher risk of recent exposure to common colds at a period when they were still prevalent (February-March). In contrast, the prepandemic samples had been collected at various times over the years but a large proportion in June 2008, some months after the cold season peak and the responses to HCoV are known to be shortlived (56). In line with this, levels of IgG to HCoV N FL decreased from M0 to M1 in seronegative and rRT-PCR negative individuals. It could also be a mixture of the two scenarios due to the wide range of antibody levels and overlap between groups. Therefore, the season of sample collection is an important variable to take into account. Consistent with crossreactive immune recognition with endemic and other emerging HCoVs, SARS-CoV-2-reactive CD4^+^ T cells have also been detected in ∼40-60% of unexposed individuals (35, 37). N is also a representative antigen for T cell response in vaccine studies, inducing SARS-CoV-specific T cell proliferation and cytotoxic activity (57, 58). However, other studies have reported a lack of N crossreactivity for antibody responses, with specificities >94%, which may be due to the protein regions included in the tests and/or the characteristics of the prepandemic controls included (59)(60). In the case of SARS-CoV-1, it has been reported that IgGs crossreact with the N antigen of endemic coronavirus, but only rarely in the other way round (42). To assess if antibodies to SARS-CoV-2 and HCoV N proteins were crossreactive and in which direction (28, 42, 61), we tested the longer-lasting IgGs responses to N FL from 229E, NL63, HKU1 and OC43. These HCoV are widely distributed, with 229E and OC43 causing 15–29% of all common colds (62). 229E, OC43 and NL63 tend to be transmitted predominantly during the winter season in temperate-climate countries, while NL63 (accounting for an estimated 4.7% of common respiratory diseases) have a Spring-Summer peak. Data on SARS-CoV-1 reported false-positive results obtained from a recombinant SARS-CoV-1 N ELISA due to the crossreactivity with antibodies to HCoV-OC43 and HCoV-229E (26, 33). Our data suggest that IgGs to SARS-CoV-2 N also recognise N229E N, and that IgGs to HKU1 N recognise SARS-CoV-2 N, but not strongly. IgG levels to NL63 N were higher in pandemic positive vs. prepandemic samples but similar for other HCoVs. Correlations of SARS-CoV-2 N CT-short with HCoV N FL were similar to those seen for N FL. When combining all HCoV N FL IgG levels, the positive correlation with SARS-CoV-2 N FL responses improved by excluding OC43. IgG responses to OC43 N correlated the least to SARS-CoV-2 in the rest of analyses, although it had a highly significant correlation with the alpha-HCoV 229E and NL63, in contrast to other reports (62). Interestingly, in rRT-PCR negative pandemic samples, there was a significant inverse correlation with IgG to OC43 HCoV that is the beta-HCoV with the highest sequence homology with SARS-CoV-2 N protein, suggesting a potential role on immunity. If preexisting IgG to OC43 bind to SARS-CoV-2, high antibody levels could neutralize SARS-CoV-2 infection upon exposure, resulting in sterile protection or low viral loads below the rRT-PCR detection threshold (rRT-PCR negative) that translates into low/no exposure and consequently less antibodies to SARS -CoV-2. If rRT-PCR individuals had not been exposed at all to SARS-CoV-2 (RBD seronegatives), then what would be measured as SARS-CoV-2 IgG might be IgG from crossreactive HCoVs. Nevertheless, a limitation of this study is that the design and sample size did not allow unraveling the potential effect of baseline HCoV antibodies on the acquisition of COVID-19 immunity (63), which will be addressed in future longitudinal follow up analysis of our cohorts. This effect could be neutral, positive (boost of responses) or negative, whereby having antibodies to certain viruses could adversely determine the profile of responses upon SARS-CoV-2 infection (‘original antigenic sin’) (44, 45, 48) or interfere by antigen masking. Findings from a recent study argue against a protective role for crossreactive HCoV T cells in SARS-CoV-2 infection (37). In our study, we speculate that preexisting OC43 HCoV IgGs could be protective and, as a result, incoming SARS-CoV-2 infections would be more controlled and thus fewer antibodies induced against them.

Unexpectedly, higher serological correlations were consistently obtained for SARS-CoV-2 and alpha-rather than beta-HCoVs despite having less homology at the primary aminoacid sequence level. The reactivity of the alpha- and beta-HCoVs clustered together within families, also by PCA analysis (data not shown). Therefore, it appears that crossreactivity is also and mostly driven by homologies at the conformational rather than the linear level.

Overall, data indicated various degrees of moderate crossreactivity for SARS-CoV-2 and HCoV N antigens, but patterns were heterogeneous and not very strong, possibly as a result of a complex polyclonal response in which different epitopes in the various viral antigens would have a wide spectrum of serological recognition and mixed binding avidities.

Our study had some limitations. We did not have access to samples from HCoV infected (rRT-PCR positive) individuals and thus the crossreactivity among the N antigens could not be fully assessed (64). In addition, we did not test IgG responses to other domains of N (like the NT that has significant sequence homologies except for a small aminoacid region) or other structural proteins (54). Our preliminary results show that crossreactivity may also exist for M and S2 (60), due to antibody recognition in some prepandemic samples, as it had been similarly described for SARS-CoV infections (65, 66). In fact, different antigens could have different levels of crossreactivity, as seen in a recent study of HCoV S antibody responses that reported an increase in IgG to OC43 (but not 229 or NL63) S proteins with SARS-CoV-2 infection (67). This observation contrasts with our data that suggested boost of N IgG from M0 to M1 for 229 and NL63 but not for OC43. Other respiratory pathogens that are common coinfections may also have crossreactivity with SARS-CoV-2 and partially explain our results (68).

In conclusion, the N protein of SARS-CoV-2 is immunogenic but it is also recognised by partially crossreactive HCoV antibodies. More specific epitopes are located in the N CT-short region and could be used for seroprevalence and diagnostics purposes. Future larger prospective studies should determine whether the N antigen is a target of protective immune responses to COVID-19 and therefore a promising vaccine candidate together with S, and the role of preexisting HCoV antibodies in acquired immunity.

## METHODS

### Antigens

#### SARS-CoV-2

Heterologously expressed N FL protein was either purchased from a commercial source (GenScript, Z03480), or purified in house after HEK cell (69) or *Escherichia coli* expression (25). Furthermore, two *E. coli* expressed His-tagged N protein constructs comprising the NT domain (residues 43-180) and CT domain (residues 250-360) were purified by affinity chromatography, as described (25). A shorter fragment of the CT region (CT-short), with a lower percentage of identity with other HCoVs and located within a putative N protein immunodominant region (EP4, residues 348-416) (24, 28), was also heterologously expressed in *E. coli* cells and His tag-purified, as described (25). Finally, an even shorter peptide, based on a less conserved region of the CT end (QRQKKQQTVTLLPAADLDDFSKQ, residues 384-406) was synthesized (CT-peptide) **(Figure S1)** (54). Additional information on the N constructs is provided in the **Supplemental material**.

#### HcoV

N FL recombinant proteins from OC43, HKU1, NL63 and 229E HCoVs were codon-optimized for *E. coli* heterologous expression and His tag-purified as in (25).

### Study volunteers and samples

We analyzed three set of samples, (i) prepandemic plasmas from healthy adults collected before the COVID-19 outbreak (negative controls, n=128), (ii) pandemic plasmas from health care personnel working at Hospital Clínic in Barcelona (Spain) collected as part of a study on SARS-CoV-2 seroprevalence at the March-April 2020 outbreak (n=578) of which SARS-CoV-2 infected cases were asymptomatic or had mild symptoms (27, 50), and (iii) pandemic plasmas from 49 COVID-19 patients recruited at the Clínica Universidad de Navarra in Pamplona (Spain), of which 47 had severe symptoms and were hospitalized and 2 had mild symptoms (25).

For the characterization of the immunogenicity of the various SARS-CoV-2 N protein constructs, we used all prepandemic and 104 pandemic samples. For the analysis of HCoV N FL cross-reactivity, we selected 30 prepandemic samples with the highest levels of IgG to SARS-CoV-2 N FL and 30 prepandemic samples with the lowest levels, among our set of negative controls tested previously (**Figure S6**) (25). Pandemic samples included: (i) 60 plasmas from individuals with a SARS-CoV-2 infection confirmed by rRT-PCR and seropositive for RBD tested in our prior studies (27, 50) (29 plasmas with the highest and 31 plasmas with the lowest levels of IgG to SARS-CoV-2 N FL), (ii) 30 plasmas from individuals with a negative rRT-PCR and RBD serology but with high IgG to N FL, and (iii) 7 negative individuals at recruitment M0 who later got infected and seroconverted for RBD at M1. For the analysis of factors associated with antibody levels to SARS-CoV-2 N FL and their kinetics, we tested all the plasma samples available from the cohort of health care workers in Barcelona at baseline (M0=578) and 1 and 3 months later (M1=565, M3=70) (27, 50).

### Study approval

The research was carried out according to the principles of the Declaration of Helsinki and informed consent was obtained. Samples analyzed in this study received ethical clearance for immunological evaluation and/or inclusion as controls in immunoassays, and the protocols and informed consent forms were approved by the Institutional Review Board at Hospital Clínic in Barcelona (Refs. CEIC-7455 and HCB/2020/0336) or Universidad de Navarra (Ref. UN/2020/067) prior to study implementation.

### Measurement of IgM, IgG and IgA antibodies

qSAT assays to measure plasma IgG, IgA and IgM against SARS-CoV-2 N proteins were performed as reported (27) and analyzed in a Luminex 100/200 instrument. For the assessement of IgG crossreactivity between HCoV and SARS-CoV-2 N proteins, we applied an optimized protocol, as described (25), and samples were analyzed with a FlexMap 3D instead of a Luminex 100/200 instrument, to increase the dynamic quantification range of antibody values in high responders (**Figure S7**). Briefly, N proteins coupled to magnetic microspheres (Luminex Corporation, Austin, USA) were incubated with plasma samples (1/500 and/or 1/3500 dilutions) or blank controls in 96-well plates. For the CT-peptide, avidin-beads were used that bound the biotin conjugated to a PEG12 biopolymer linked to the peptide, and the coupling performed following manufacturer’s instructions. Before multiplexing protein-coupled MagPlex beads, we tested for potential interference among N constructs comparing to singleplex assays (**Figure S7**). After sample incubation with beads, plates were washed and a labeled secondary antibody (anti-human IgG, IgM, or IgA) was added. Following the last incubation, plates were washed and read in a Luminex xMAP® analyzer. Crude median fluorescent intensities (MFI) and background fluorescence from blank wells were exported using the xPONENT software.

### Statistics

Boxplots and Wilcoxon Rank Sum tests were used to compare antibody levels to each N protein between study groups. To assess for crossreactivity, we performed correlations (Spearman) and heatmaps of antibody responses to SARS-CoV-2 and HCoV N antigens in prepandemic and pandemic samples separately, stratifying by rRT-PCR positivity and by high vs. low anti-N FL IgG responders, as appropriate. Seropositivity was defined by a cutoff calculated with prepandemic samples as 10 to the mean plus 3 standard deviations of log_10_-transformed MFI. A variable called magnitude of responses to HCoV N FL was created by adding up the levels of IgG to the four HCoV N FL. To evaluate the factors associated with levels of antibodies to SARS-CoV-2 N FL, we used Wilcoxon Rank Sum test. The LOESS (locally estimated scatterplot smoothing) method was used to fit a curve to depict kinetics of antibody levels over time. The analysis was carried out using the statistical software R studio version R-4.0.2 (70) (packages used: tidyverse (71), pheatmap (72) and corrplot (73)).

## Data Availability

Data are available upon request to the corresponding authors

## Author contributions

CD, GM, RA and LI designed the serological study; RS analyzed the data; AJ and MV performed the Luminex experiment; JC, NRM, MP, CC and LI designed and/or produced the recombinant proteins; RLA, LFB and AT performed the peptide studies; MT, FCT, GR, AM, GM, CD and AGB recruited, followed up and sampled human volunteers; AM and CC contributed to rRT-PCR determinations; CD wrote the first manuscript; all authors reviewed and approved the manuscript.

## Acknowledgements

We thank the volunteers who donated blood for COVID-19 studies and the clinical and laboratory staff who participated in the sample collection and processing. Special thanks to ISGlobal lab colleagues D. Barrios, P. Cisteró, L. Puyol, R.A. Mitchell, C. Jairoce, S. Alonso, J. Moreno, and those involved in data management and/or recruitment of volunteers and patients at the Pamplona and Barcelona hospitals, J.L. del Pozo, M. Fernández-Alonso, S. Sanz, S. Méndez, A. Llupià, E. Chóliz, A. Cruz, A. Figueroa-Romero, S. Folchs, M. Lamoglia, N. Ortega, N. Pey, M. Ribes, N. Rosell, P. Sotomayor, S. Torres, S. Williams, S. Barroso, A. Trilla and P. Varela.

