## Supplemental material for "Immunogenicity and crossreactivity of antibodies to SARS-CoV-2 nucleocapsid protein"

**Supplemental Methods and Results**

**N proteins and peptide constructs**

*Expression and purification of recombinant proteins in heterologous systems.* Full-length (FL) N proteins were expressed in freestyle HEK F cells with a C-terminal (CT) strep tag, and purified as described (69). Likewise, an *E. coli* optimized version of N was cloned in a pET pET22b vector, heterologously expressed in *E. coli* BL21 DE3 cells and His-tag purified, as described elsewhere (25). The constructs were compared for Ig quantification to a commercial N FL protein expressed in *E. coli* with an N-terminal (NT) His tag (Z03480, GenScript). NT domain (residues 43-180) and CT domain (residues 250-360) constructs, considering secondary structure predictions, were cloned into a pETM14 plasmid with NT 6xHis-tag, transformed into *E. coli* BL21 DE3 cells, induced with IPTG and purified using HisTrap Ni-NTA columns (25).

Considering the high similarity between the N proteins of SARS-CoV and SARS-CoV-2 (>90% pairwise-identity) (**Figure S1A**) a shorter fragment of the CT region with a lower percentage of amino acid identiy with other HCoV (CT-short), located within an immunodominant region (EP4) of N (24, 28), was cloned in a plasmid, transformed into *E. coli* BL21 DE3 cells, expressed and His-tag purified. **Figure S1B** depicts a diagram describing all the different fragments expressed, purified and used for Ig analyses in plasma samples.

*E. coli* codon optimized versions of OC43, HKU1, NL63 and 229E N FL proteins were cloned in pET22b expression vectors, fusing a CT 6xHis-tag, transformed in *E. coli* BL21 DE3, induced with IPTG and purified by affinity chromatography using HisTrap columns. Purity and integrity of all purified recombinant FL proteins and fragments was checked by Coomassie stained SDS-PAGE, showing >90% purity. A layout of the HCoV N proteins and the % pairwise-identity with previously described SARS antigenic regions (24, 28) can be found in **Figure S1B**.

*N CT-peptide*. A peptide based on SARS-CoV-2 N CT residues 384-406, was synthesized and chemically modified at BCNPeptides (Barcelona, Spain) with a PEG12 biopolymer, conjugated to biotin, and purified by High Pressure Liquid Cromatography (HPLC). Modifications were done to avoid steric problems related to small size, for an enhanced performance in Luminex assays.

**Testing of additional nucleocapsid (N) protein constructs**

To complement the characterization of the N FL construct produced in-house at ISGlobal, we compared antibody levels in prepandemic and pandemic plasma samples to those obtained with a recombinant protein from a commercial source (GenScript). The correlation of antibody responses between the two antigens was very significant (rho=0.77-0.96) (**Figure S3A**), indicating that the non-specific responses observed in prepandemic samples were not due to a suboptimal quality of the purified proteins. In fact, the produced antigens were of high purity (> 90%). We also compared the antibody responses between an *E. coli* N FL construct and an N FL protein produced in eukaryotic cells at CRG to test whether non-specific responses were due to the expression in the prokaryotic system. The antibody recognition was very similar in the two expression systems (rho=0.62-0.81) (**Figure S3B**) and therefore we concluded that our N FL construct accurately represented the N protein in its native form on the SARS-CoV-2 virus and that the responses in negative controls were likely due to crossreactivity with other coronaviruses, which we investigated subsequently.

We compared the seroreactivity of two SARS-CoV-2 N proteins (N FL and N CT-short) with four possibly crossreactive N FL proteins from 229E, NL63, HKU1 and OC43 HCoV against a pool of COVID-19 positive samples. The two SARS-CoV-2 N proteins showed the highest levels of IgG responses, followed by the N FL from the two alphacoronaviruses, and finally the N FL from the two betacoronaviruses, which were also recognized by the COVID-19 positive pool (**Figure S4**).

**Supplemental Figures**

**Figure S1.**

**A.** SARS-CoV and SARS-CoV-2 N pairwise alignment. Highlighted sequences indicate the four immunodominant (EP1 to EP4) regions previously identified (24), which show a high percentage of aminoacid pairwise identity (EP1-EP3 > 90% & EP4 84.1%).

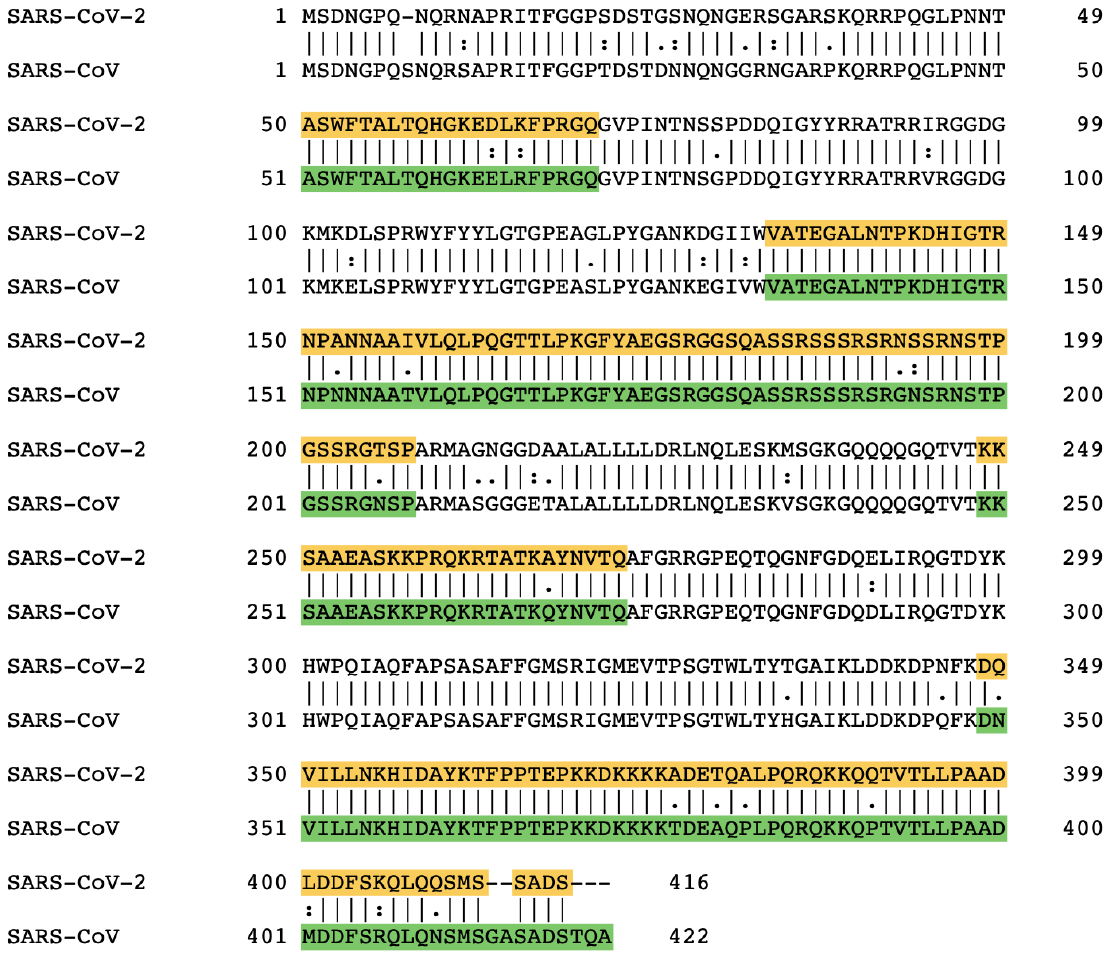

**B**. Diagram depicting SARS-CoV-2 and HCoV N proteins (28). The EP immunogenic regions (24) are marked for every protein, including the percentage (%) of amino acid (AA) pairwise identity. Heatmap color scale indicates lesser to higher % pairwise identity from red to green, respectively. Blue bars below show the different protein fragments used in this study.

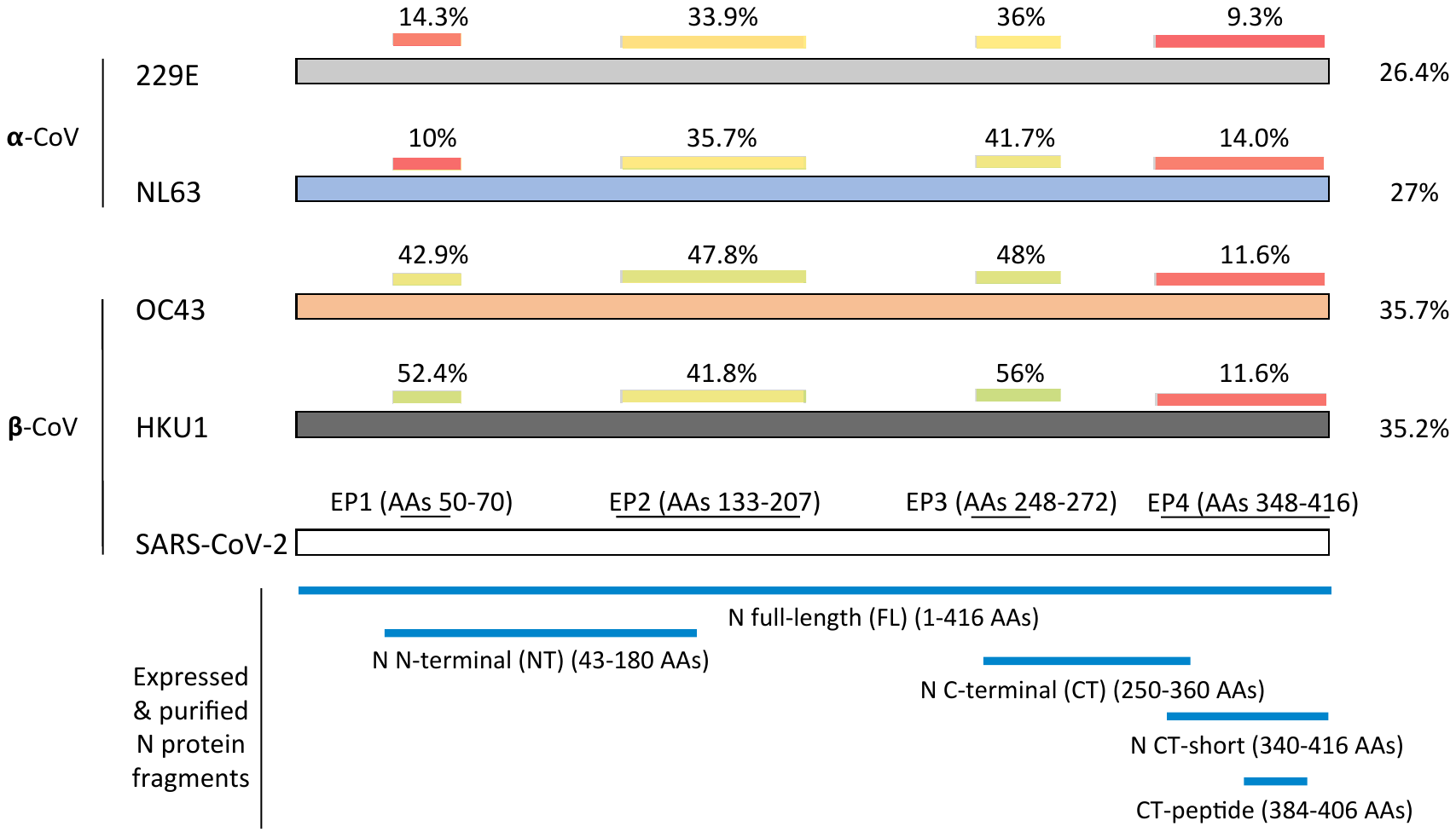

**Figure S2.** Levels (median fluorescence intensity, MFI) and seropositivity of antibody responses to nucleocapsid (N) SARS-CoV-2 proteins in prepandemic and pandemic individuals. **A)** Antibodies to N FL in plasma (1/500 dilution) in a seroprevalence survey in a cohort of 565 health care workers one month after the March-April 2020 first wave (50). **B.** Effect of plasma dilution on seropositivity estimates in 104 COVID-19 pandemic (black) and 128 prepandemic (grey) samples (25). Reactivity of prepandemic plasmas and cutoffs decreased in 1/3500 compared to 1/500 dilution depending on the Ig isotype and antigen. Cutoff values, indicated by dashed lines, are calculated based on the mean plus 3 standard deviations of 47 (A) or 128 (B) prepandemic controls. Antigens: N full-length (FL) from ISGlobal, N-terminus (NT) and C-terminus (CT) domains from CRG.

**A.**

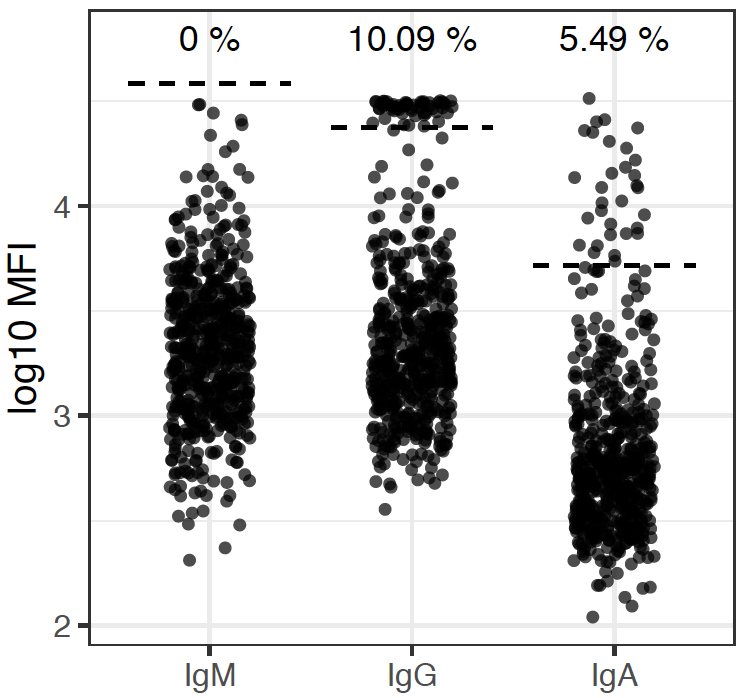

**B**.

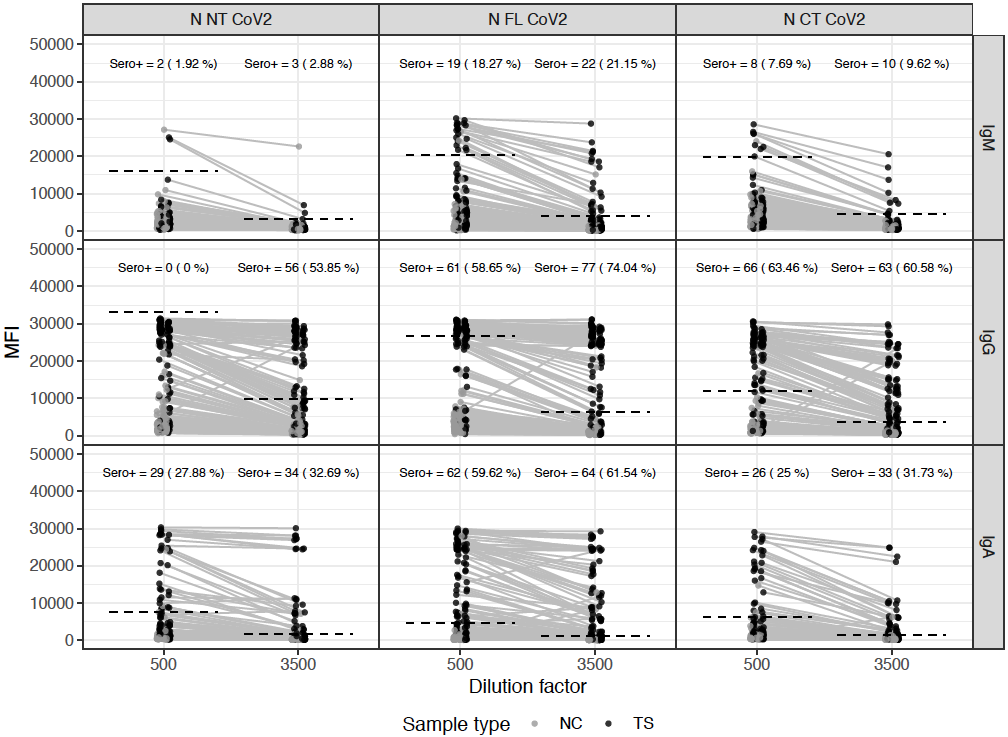

**Figure S3.** Correlations of antibody levels (log_10_ MFI) among different nucleocapsid (N) full-length (FL) constructs of SARS-CoV-2 in pandemic and prepandemic plasma samples. **A)** N FL purified in-house at ISGlobal compared to commercially purchased N FL (GenScript), both expressed in *Escherichia coli.* **B)** *E. coli*-expressed N FL produced in-house at ISGlobal compared to HEK F mammalian cells-expressed N FL produced in-house at CRG. Rho and p-values (p) are calculated by Spearman, shaded areas represent 0.95 confidence intervals

**A.**

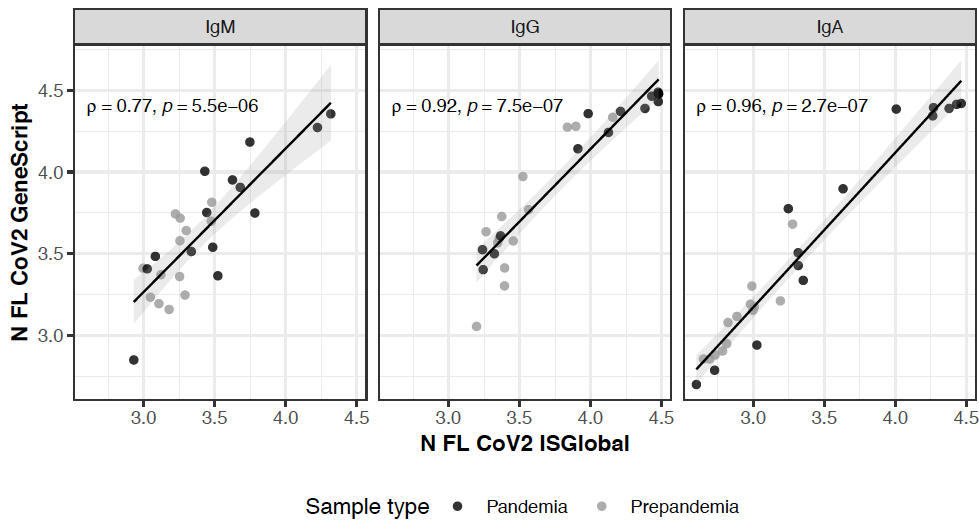

**B.**

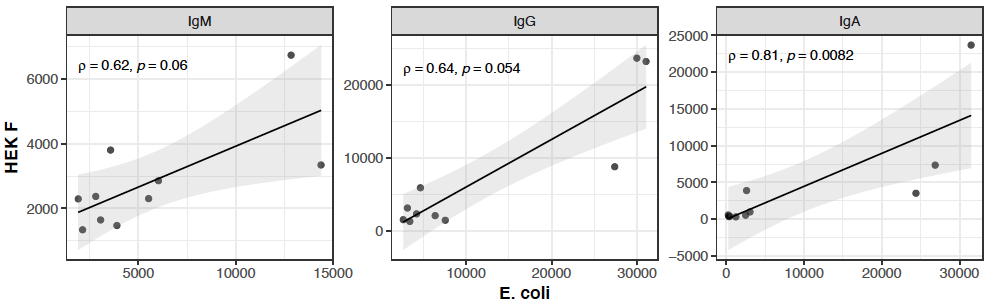

**Figure S4. Titration curves of IgG to the nucleocapsid (N) constructs from SARS-CoV-2 and HCoV against a pool of plasmas from pandemic COVID-19 cases.** IgG levels, expressed as mean fluorescence intensity (MFI), were higher for SARS-CoV-2 (more prominent in full length [FL] vs C-terminus [CT] short) than for the HCoV N FL constructs. However, recognition of HCoV N proteins is substantial (alpha-coronaviruses > beta-coronaviruses), indicating both specific and crossreactive responses being measured.

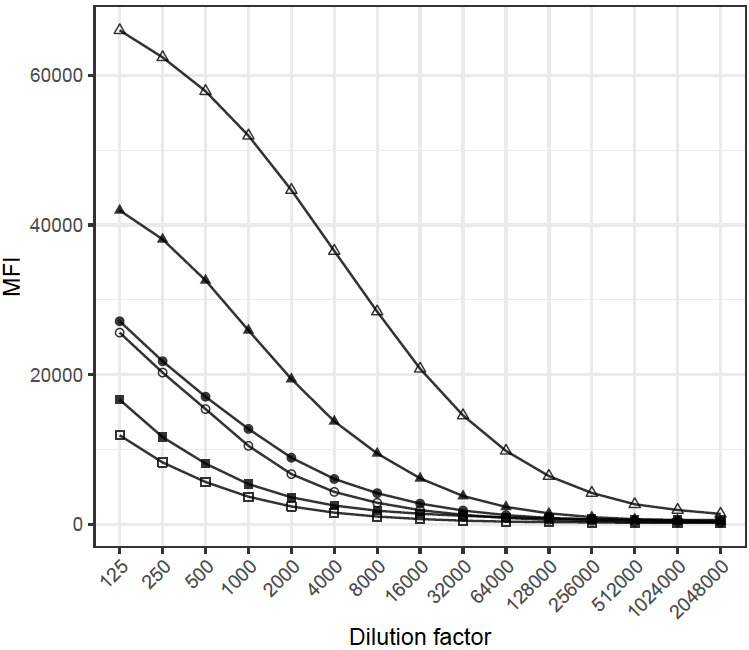

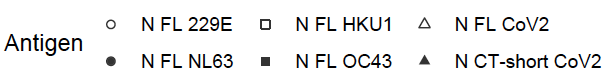

**Figure S5. Relationship between IgG responses to N proteins of SARS-CoV-2 and HCoV in prepandemic and pandemic samples**. **A)** Correlations between IgG levels to the different N constructs. The intensity of the red-yellow color scale corresponds to the intensity of the rho Spearman coefficient. Significance of p values is indicated by: * p<0.05, ** p<0.01, *** p<0.001. **B)** Heatmaps of IgG responses to N constructs by individual. The intensity of the red-yellow color scale corresponds to the intensity of the IgG MFIs.

**A**

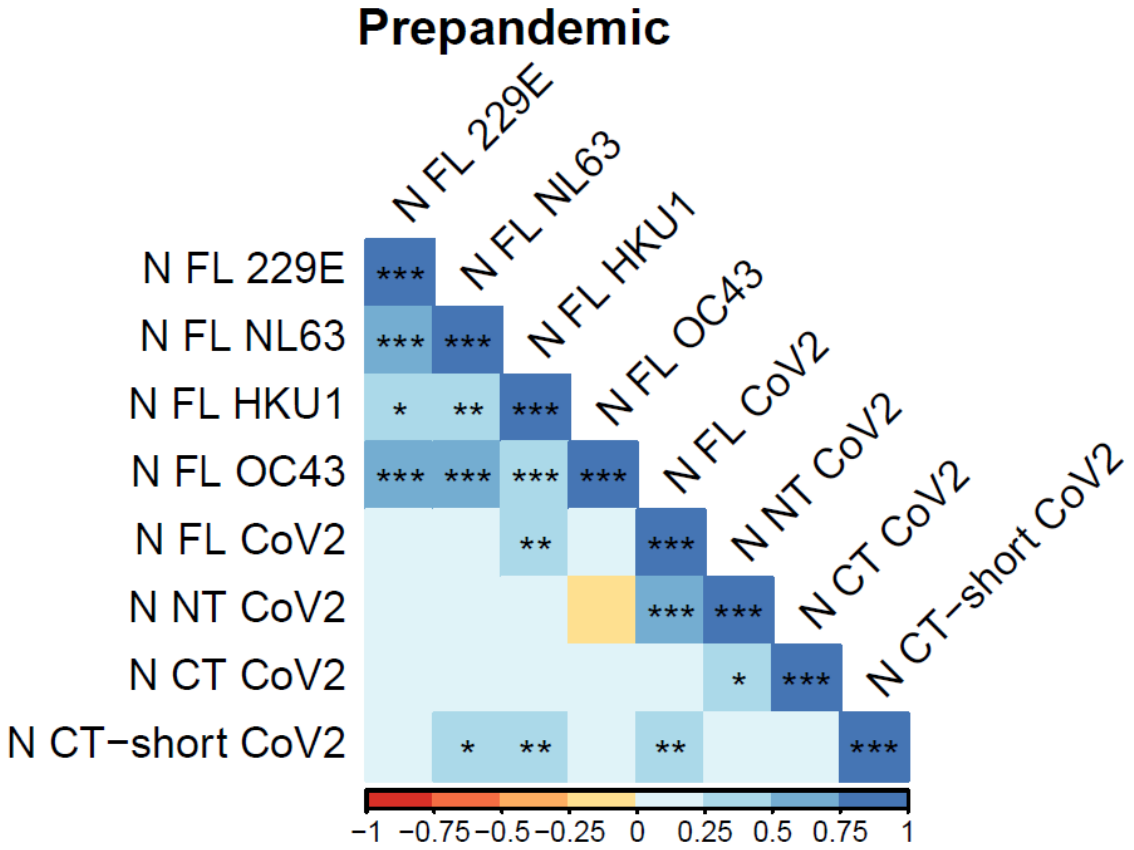

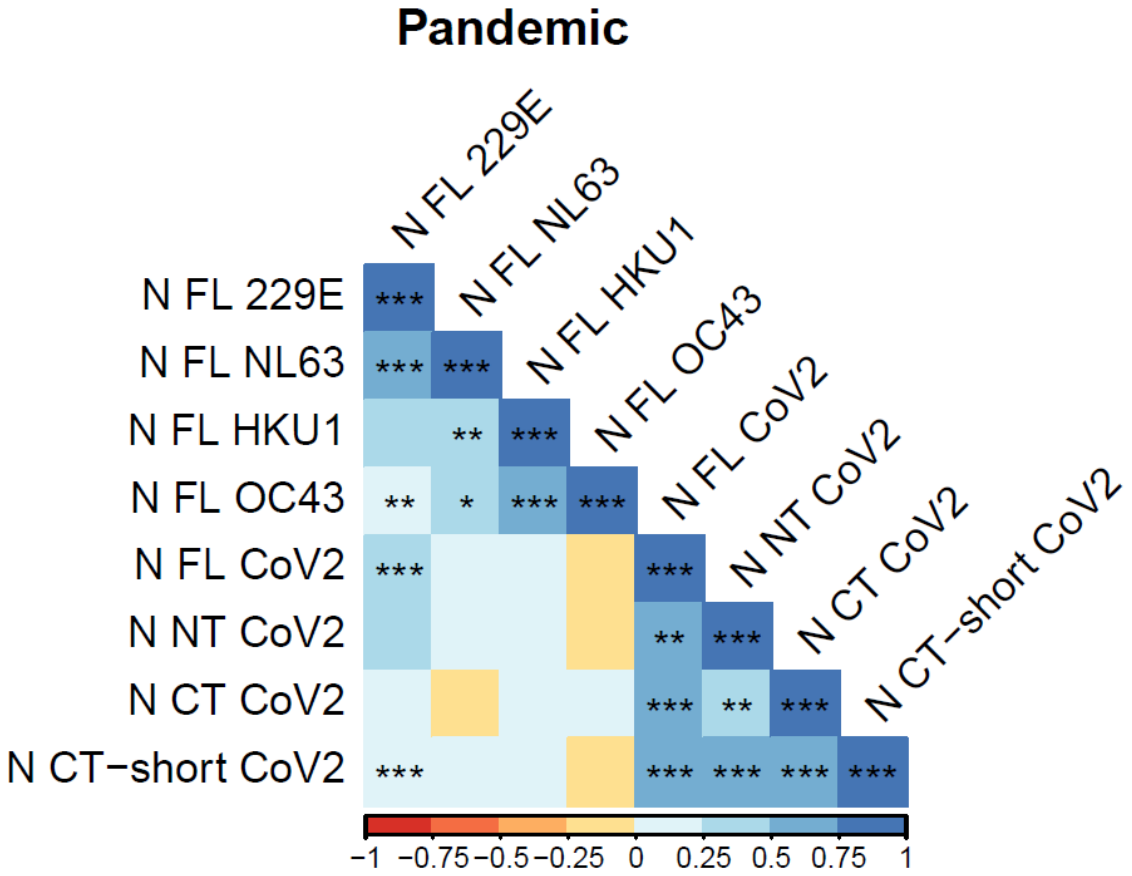

**B**

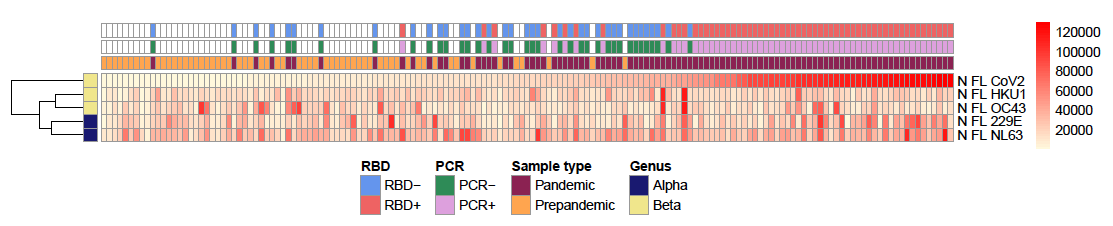

**Figure S6.** **Study groups for the crossreactivity analysis of IgG to SARS-CoV-2 and HCoV nucleocapsid (N) protein.** Antibody responses to HCoV N FL were tested in groups of prepandemic (n=60) and pandemic (n=60, rRT-PCR positive or negative, RBD IgG seropositive or seronegative) samples classified according to the levels of IgG response to SARS-CoV-2 N FL (high vs. low) obtained in a previous study (25), and which levels are shown here. Pandemic samples included 29 plasmas (12 month [M]0, 17 M1) with the highest and 31 plasmas (12 M0, 19 M1) with the lowest levels of IgG to SARS-CoV-2 N FL, and 30 plasmas (22 M0, 8 M1) from individuals with a negative rRT-PCR and RBD serology but with high IgG to N FL, as well as 7 negative individuals at M0 who later got infected and seroconverted for RBD at M1. The lower and upper hinges of the boxplots correspond to the 25^th^ and 75^th^ percentiles (IQR) and extend 1.5 * IQR from the hinge. N FL+: high IgG responder; N FL-: low IgG responder.

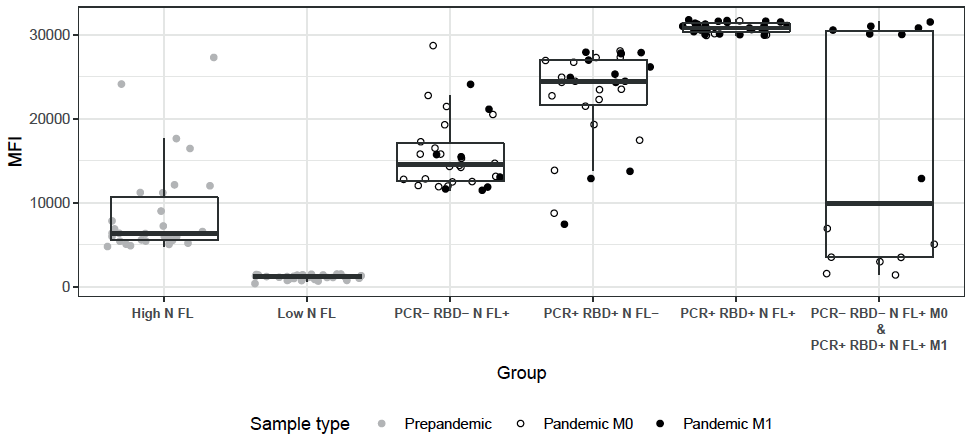

**Figure S7. Comparison of antibody responses to nucleocapsid (N) proteins of SARS-CoV-2 and HCoV measured in singleplex (SP) or multiplex (MP) in two Luminex instruments.** Serial dilutions of a positive control measured against HCoV and SARS-CoV-2 antigens in a Luminex 100/200 (LM) or FlexMap3D (FM) instrument in both singleplex (SP) and multiplex (MP). No interference was observed in MP. FP showed a higher dynamic range. BL, blank. MFI: median fluorescence intensity (antibody levels).

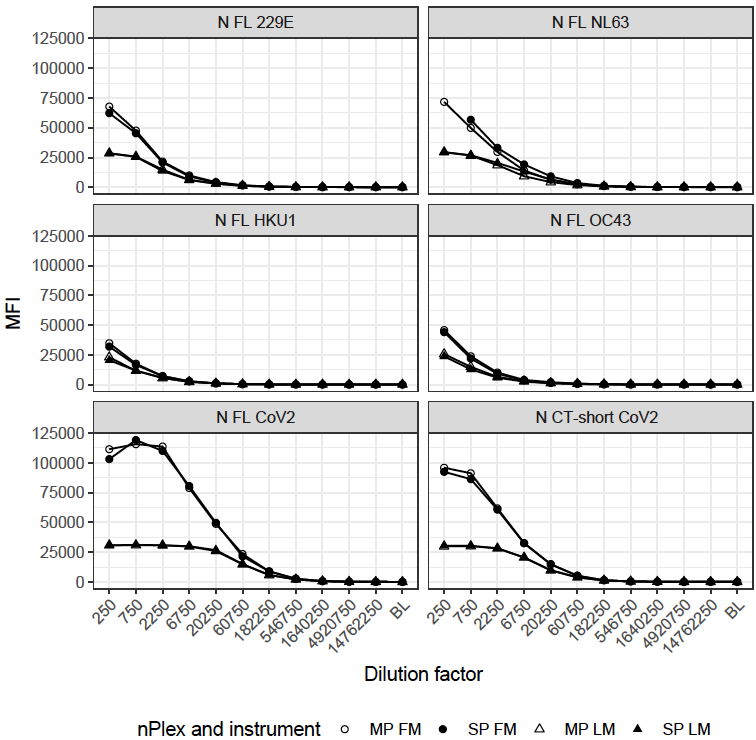
